# Effects of Prophylactic Low-Dose Acetazolamide on the Chemoreflex Control of Breathing and Acute Mountain Sickness at High Altitude

**DOI:** 10.64898/2026.07.29.26359164

**Authors:** Abel Vargas, Hannah Pagliero, Veronica L. Penuelas, Herlinda Bergman, Donna Amaya, Samantha Limvalencia, Erica C. Heinrich

## Abstract

**Introduction:** High altitude is a physiologically stressful environment due to low barometric pressure and low oxygen availability. This hypoxic stress commonly leads to Acute Mountain Sickness (AMS) during high altitude travel. Additionally, among the physiological adaptations to sustained hypobaric hypoxia, ventilatory acclimatization is characterized by increases in the ventilatory chemoreflex responses to hypoxia and hypercapnia. This study aimed to determine how low-dose prophylactic use of Acetazolamide (ACZ), as utilized for recreational high-altitude travel, impacts these breathing reflexes.

**Methods:** In a double-blind, randomized, placebo-controlled design, participants received 125 mg of ACZ taken twice daily (N=9) or placebo (N=9) starting 2 days prior to ascent to high-altitude (3801 m) and continued until departure. Ventilatory chemoreflex tests were performed at sea level prior to treatment and after 2 days at high altitude.

**Results:** ACZ reduced AMS severity on the first two days at high altitude. ACZ also lowered end-tidal P_CO2_, indicating elevated baseline alveolar ventilation. Among the ventilatory chemoreflex characteristics, the ventilatory recruitment threshold was significantly lower at high altitude in both groups. We also found a highly significant three-way interaction between location, treatment, and P_CO2_ on the hypoxic ventilatory response (HVR), indicating that the relationship between P_CO2_ and the amplitude of the reflex increase in breathing in response to hypoxia is impacted by ACZ.

**Conclusions:** ACZ may sensitize the interaction between the peripheral and central chemoreflex responses. This data provides insight into the mechanisms underlying ACZ’s beneficial effects on sleep quality and AMS symptoms at high altitude.

**Trial registry:** ClinicalTrials.gov

**Registration number:** NCT07517068

## INTRODUCTION

High altitude (HA) is a physiologically stressful environment due to low barometric pressure resulting in low oxygen availability. As a result, acute exposure to this environment can cause Acute Mountain Sickness (AMS), characterized by headache, nausea, lightheadedness, and often coupled with poor sleep quality.^1^ Acetazolamide (ACZ) is the best prophylactic treatment for prevention of AMS and more severe conditions, including cerebral and pulmonary edema during HA travel. ACZ is a carbonic anhydrase inhibitor which reduces renal absorption of bicarbonate and increases bicarbonate diuresis, causing a metabolic acidosis that results in higher arterial oxygen levels by stimulating increased minute ventilation.^2^

The neural control of breathing, both baseline ventilation and chemoreflex sensitivity to oxygen (O_2_) and carbon dioxide (CO_2_), are significantly altered during acclimatization to hypoxia.^3–5^ The amplitude of the hypoxic and hypercapnic ventilatory responses (HVR, HCVR, respectively) has been shown to increase progressively over the first several days at altitude.^6^ Over the past several decades, multiple methods have been used to quantify these chemoreflex responses. One major difference between methods is whether or not the hypoxic or hypercapnic challenge is maintained at a constant level over a period of several minutes (steady-state) or if O_2_ and CO_2_ levels are allowed to change progressively over time, such as in rebreathing methods.^7–9^ One benefit of a particular rebreathing method described by Duffin (2007) is the ability to identify the ventilatory recruitment threshold (VRT), or the CO_2_ partial pressure (P_CO2_) at which the linear increase in minute ventilation in response to increased CO_2_ begins (**Figure 1**).^8^ Steady-state methods may miss the VRT since as few as two discrete O_2_ and CO_2_ levels are chosen, and a linear fit between minute ventilation and O_2_ or CO_2_ is assumed. Another possible benefit of rebreathing methods is that they allow the quantification of the interaction between the peripheral and central components of the ventilatory chemoreflex.^10^

**Figure 1.**
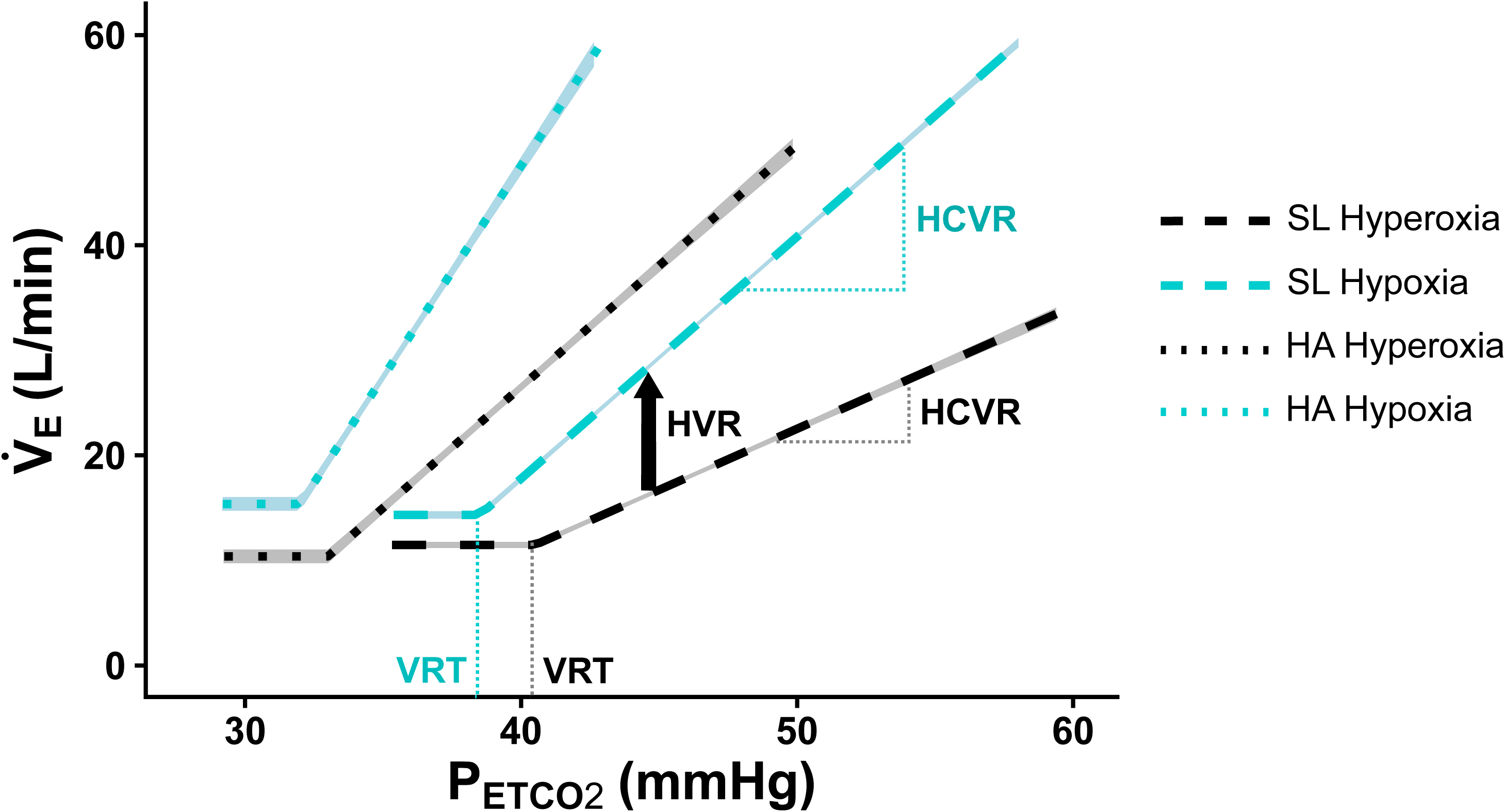
Representative trace of chemoreflex test results from one participant. Average minute ventilation (V_E_) is plotted as a function of end-tidal P_CO2_ (P_ETCO2_) at sea level and high altitude under hyperoxic (inspired PO_2_ = 228 mmHg, equivalent to 30% at SL) and hypoxic (end-tidal PO_2_ = 50 mmHg) conditions. Hyperoxic tests effectively reduce or silence input from peripheral chemoreceptors to provide the central chemoreceptor response to CO_2_ alone, while the hypoxic tests include stimulation of both central and peripheral chemoreceptor responses to CO_2_ and O_2_. 95% confidence intervals for the lines of best fit are highlighted. Components of the chemoreflex responses to hypoxia and hypercapnia are labeled for the sea level tests only. HVR: hypoxic ventilatory response; HCVR: hypercapnic ventilatory response; VRT: ventilatory recruitment threshold.

While prior work has evaluated the impact of ACZ on ventilatory control at sea level and high altitude, some gaps in knowledge remain. Prior studies typically utilized 250 mg taken twice per day, a larger dose that may provide higher power to detect treatment effects.^11–14^ However, in recent years, the preventative dose has been recommended as 125 mg taken twice per day, and doses as small as 62.5 mg twice per day have been shown to be effective at reducing AMS symptoms.^15,16^ Furthermore, many studies report no impact of ACZ on the ventilatory sensitivity to changes in PO_2_ and P_CO2_^11,17,18^ despite a significant increase in baseline minute ventilation and associated improvements in oxygenation parameters and AMS symptoms.^11,17^ This study therefore aimed to determine if the lower ACZ dose, which is effective at mitigating AMS symptoms, has similar impacts on the control of breathing after two days of acclimatization. We also aimed to thoroughly characterize any effects of ACZ on changes in central and peripheral chemoreflex sensitivities, as well as the VRT, which may have been missed in prior studies using steady-state methods. We hypothesized that this lower dose would still significantly elevates baseline minute ventilation and produces a leftward shift in the HCVR, indicating enhanced ventilatory sensitivity to CO_2_, without impacting the isocapnic HVR, as previously reported for higher dosages.

## METHODS

### Ethical approval

This study was approved by the University of California Riverside Institutional Review Board (protocol #30687). The study was conducted in accordance with the *Declaration of Helsinki*, including registration in a database (clinicaltrials.gov NCT07517068). A digital version of the complete consent form was provided to all participants prior to their first study visit to review on their own. During their first study visit, we reviewed the written consent form with participants, provided them with time to ask any questions, and each participant provided written, informed consent in their native language (English). All experiments were conducted by trained and qualified personnel and with oversight by a licensed medical doctor.

### Participant demographics

Healthy men and women were recruited. Exclusion criteria included age less than 18 or greater than 65, history of significant cardiovascular or pulmonary disease including sleep disordered breathing, prior history of high altitude pulmonary or cerebral edema, use of interfering medications, current or suspected pregnancy, travel above 8,000 m elevation within 30 days of the first measurements, smoking (including regular use of vapes or marijuana), and current or recently treated systemic or serious local infection. Participants were asked to refrain from consuming alcohol or caffeine and to avoid any anti-inflammatory medications starting 12 hours before their first appointment and throughout the study.

Nineteen participants were recruited; however, one participant became ill after completing baseline sea level measures, did not travel to high altitude, and was excluded. A second participant did not complete chemoreflex testing at high altitude due to unexpected visual symptoms following hyperventilation, leading to premature termination of the test, but their baseline data was included in the analyses. **Table 1** provides a detailed breakdown of participant demographics.

**Table 1.**
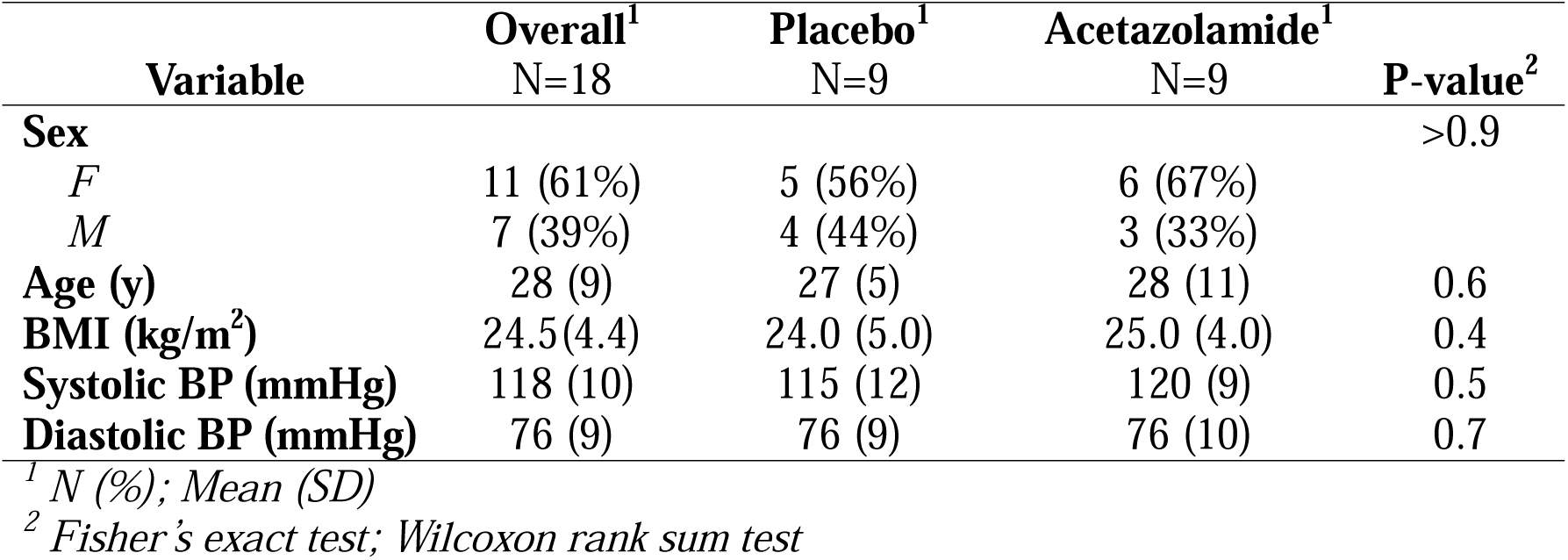
Participant demographic information.

| **Variable** | **Overall<sup>1</sup>**<br>N=18 | **Placebo<sup>1</sup>**<br>N=9 | **Acetazolamide<sup>1</sup>**<br>N=9 | **P-value<sup>2</sup>** |
| --- | --- | --- | --- | --- |
| **Sex** |  |  |  | >0.9 |
| *F* | 11 (61%) | 5 (56%) | 6 (67%) |  |
| *M* | 7 (39%) | 4 (44%) | 3 (33%) |  |
| **Age (y)** | 28 (9) | 27 (5) | 28 (11) | 0.6 |
| **BMI (kg/m<sup>2</sup>)** | 24.5(4.4) | 24.0 (5.0) | 25.0 (4.0) | 0.4 |
| **Systolic BP (mmHg)** | 118 (10) | 115 (12) | 120 (9) | 0.5 |
| **Diastolic BP (mmHg)** | 76 (9) | 76 (9) | 76 (10) | 0.7 |

<sup>1</sup> *N (%)*; *Mean (SD)*
<sup>2</sup> *Fisher’s exact test*; *Wilcoxon rank sum test*

### Experimental overview

The data reported here was collected as part of a larger study with multiple experimental aims but represent an independent experiment with its own a-priori hypothesis. This study was conducted in a placebo-controlled, double-blind, randomized cohort design. Participants were prospectively assigned to receive either Acetazolamide (125 mg dose taken orally 2 times per day, as recommended for prophylactic prevention of AMS) or placebo (visually identical and taken 2 times per day). Participants began taking their assigned treatment starting 2 days before the day of ascent to high altitude. 2 individuals began taking their treatment 1 day before ascent due to logistical constraints in completing baseline measurements (one assigned to placebo and one to ACZ treatment). Treatment continued throughout three days at high altitude (HA1-HA3) until the morning of departure.

Participants first completed a laboratory visit at the University of California, Riverside (approximately 400 m elevation) for consenting procedures and baseline physiological measures. At this visit, participants conducted consent procedures followed by measurements of height, weight, blood pressure, heart rate, pulse oximetry, and lung function. They then completed a series of questionnaires related to their medical history and current symptoms. Participants then completed ventilatory chemoreflex testing procedures followed by consultation with a medical doctor for prescription of their assigned treatment.

Participants returned in the early morning on the day of ascent. Blood pressure, heart rate, pulse oximetry, and symptom questionnaires were completed. The group then traveled by car to Barcroft Field Station in the White Mountain Research Center (Bishop, California, USA, 3801 m elevation). The ascent profile includes a 5-hour slow ascent from 400 to approximately 1225 m elevation (Big Pine, California, USA), followed by a 1.5-2 hour ascent from 1225 m to 3801 m elevation.

While at high altitude, blood pressure, heart rate, SpO_2_, and symptoms were assessed each morning immediately following waking and in the evening. Ventilatory chemoreflex testing was performed throughout the day on the second day at high altitude.

### Physiological data collection

Resting blood pressure was collected using a manual sphygmomanometer using standard procedures. Resting heart rate and pulse oximetry were obtained with a Nellcor N600x pulse oximeter (Medtronic, Minneapolis, MN, USA) using a finger probe while the participant was seated upright with their legs uncrossed and instructed to relax. Measures were taken when values stabilized after approximately 1 minute. Lung function was measured by spirometry using an ADInstruments spirometer (model FE141) and respiratory flow head (model MLT1000L) (AD Instruments, Colorado Springs, CO, USA). Participants took three normal tidal breaths followed by one maximal expiration and a rapid, forceful 6 second expiration. This was repeated 3 times and mean values were manually collected in LabChart (ADInstruments) for forced vital capacity (FVC) and forced expired volume in 1 second (FEV_1_).

### Questionnaires

All questionnaires were completed independently by participants on a desktop computer or tablet. On the first morning at HA, questionnaires were completed on paper due to a technical issue. Each morning, participants completed the 2018 Lake Louise Acute Mountain Sickness scale.^19^ During the first session, participants were provided with instructions, and a research team member was available to answer any questions throughout the session.

### Ventilatory chemoreflex testing

#### Rebreathing protocol

HVR and HCVR measurements were collected as described previously by our group^20^, using Duffin’s modified rebreathing procedure^8^, with minor changes described below. Tests took place in a quiet, isolated room. Participants were seated in a semi-recumbent position and fitted with a three-lead electrocardiogram (Bioamp, ADInstruments) and finger pulse oximeter probe (Nellcor N-600x, Medtronic). Participants breathed through a mouthpiece connected to a system containing, in series, a respiratory filter, flow head (MLT1000L, ADInstruments), and a three-way directional valve (R2100, Vacumetrics, Ventura, CA, USA). Participants first breathed room air for five minutes. This provided us with resting measures and allowed the participants to acclimate to the mouthpiece. Participants were then asked to voluntarily hyperventilate using slow, deep, and deliberate breaths until the end-tidal P_CO2_ reached approximately 22 mmHg, which typically took 1-3 minutes. Boulet et al. (2016) show no change in ventilatory chemoreflex characteristics measured with this method across hyperventilation periods of 1, 3, or 5 minutes.^21^ Following hyperventilation, participants were quickly switched to breathing from a rebreathing bag. Once switched onto the bag, the gas partial pressures in the lungs and bag were equilibrated through 2-3 deep breaths. Participants were then asked to relax. Rebreathing continued until end-tidal P_CO2_ reached 60 mmHg, minute ventilation reached 100 L/min, or if participants indicated they were unable to continue.

Participants completed this rebreathing procedure twice. Once while maintaining hyperoxic conditions and once with hypoxic conditions. During hyperoxic test conditions, the peripheral chemoreflex response is minimized or eliminated. The hypoxic test then allows measurement of the summed peripheral and central chemoreflex responses.^10^ Thus, the HVR can be evaluated by subtracting the ventilation rate between these two response curves at any given P_CO2_ can provide the HVR. At the start of each test, the 6 L rebreathing bag contained either a hyperoxic (30% O_2_, 6.5-7% CO_2_, N_2_ balanced sea-level equivalent pressures at high altitude) or hypoxic (8.5% O_2_, 7.5-8% CO_2_, N_2_ balanced) gas mixture. End-tidal CO_2_ and inspired O_2_ measurements were collected between the mouthpiece and rebreathing bag by subsampling and analysis by an infrared CO_2_ analyzer (model 17630, Vacumetrics) and a paramagnetic O_2_ analyzer (model 17625, Vacumetrics). A small input valve at the bottom of the bag allowed oxygen to be added to the bag via a 5L oxygen concentrator (525KS, DeVilbiss Healthcare, Port Washington, NY, USA) throughout the test to maintain the target isoxic test conditions (30% inspired O_2_, or end-tidal P_O2_ of 50 mmHg). The 30% sea-level equivalent inspired O_2_ (P_O2_ = 228 mmHg in both locations) hyperoxia target was chosen to maintain an P_aO2_ of approximately 150 mmHg, providing a high enough O_2_ stimulus to minimize or eliminate peripheral drive without providing any potential hyperoxia-induced increases in ventilation^22^, although the impact of such a stimulus is debated.^23^ The end-tidal target of 50 mmHg P_O2_ was chosen because it produced a substantial desaturation within safe limits, allowed most participants to reach an end-tidal P_CO2_ of 60 mmHg at SL, and was consistent with prior studies.^24^

#### Data collection and analysis

Analog ECG, flow, and SpO_2_ data signals were collected by an 8-channel PowerLab data acquisition system (ADInstruments) and digital signals were sent to a desktop PC, monitored, and recorded in LabChart 9 software (ADInstruments) with as sampling rate of 1000/sec.

Inspiratory volume was determined by calculating the integral of the respiratory flow rates, and volumes were BTPS corrected. Resting ventilation values were determined during the last 1 minute of the 5-minute room-air breathing period at the start of each test. Data selected for rebreathing chemoreflex analyses included any data recorded after the equilibration point in the P_CO2_ channel, indicated by a temporary plateau in P_CO2_, typically occurring 4-5 breaths into the rebreathing portion of the test. Labchart data was exported to a csv format, downsampled by 1000/sec, and analyzed downstream in RStudio (RStudio, Boston, MA, USA) with R version 4.4.0. R packages *mcp* and JAGS were used to identify the VRT and HCVR slopes under hyperoxic and hypoxic conditions based on the best-fit line for minute ventilation data plotted as a function of end-tidal P_CO2_, with a single break point. The HVR was calculated as the change in minute ventilation at a single P_CO2_ level (45, 50, or 55 mmHg) across hyperoxic and hypoxic test conditions. We also calculated HVR as described by Duffin (2007) by determining the change in HCVR slope and VRT across oxygen levels.^8^

### Statistical analysis

All statistical analyses were performed in R Studio (2026.01.1) using R version 4.3.3.^25^ All variables were checked group-wise (within treatment and location) to determine if they met the assumptions of a mixed ANOVA using Shapiro-Wilks tests for normality and Levene tests for homogeneity of variance. However, assumptions of normality and homogeneity of variance were not met in several groups, so linear mixed-effect models were employed for all analyses.

Linear mixed-effects models were conducted using the *lmer* function within the *lme4* package.^26^ Each baseline respiratory variable, as well as the VRT and HCVR, was measured twice at each location (before or during a hyperoxic and hypoxic rebreathing test). Therefore, as fixed effects, we entered the full three-way interaction of location (SL, HA), Treatment (ACZ, placebo), and test (hyperoxia, hypoxia), including all main effects and two-way interactions. To account for the repeated-measures structure of the data, we specified a random-effects structure that included random intercepts for each participant, as well as random slopes for location and test by participant. This allowed the effects of location and test to vary across individual participants while estimating the correlations between the random intercepts and slopes. Since HVR measures are inclusive of hypoxic and hyperoxic tests, the variable “test” was removed from the model design for analyses of HVR.

When significant main effects or interactions were identified, we then ran post-hoc pair-wise comparisons with Tukey’s HSD p-value adjustments across the significant grouping factor using the *emmeans* function from the *emmeans* package and applying the *marginal* function to look at specific comparisons of treatments within location groups.^27^ Since p-value correction of post-hoc comparisons may be too strict for this study design, we report all p-values less than 0.1 throughout the manuscript and highlight possible trends in the event that significant effects are missing effects due to type II error.

As an alternative analysis, we also calculated changes in baseline physiological variables at HA compared to SL and conducted unpaired t-tests or Wilcoxon signed-rank tests, depending on data normality and homogeneity of variance, to determine if these delta values differed across treatment groups.

Throughout the paper and figures, p-values less than 0.05 are considered significant and p-values between 0.1 and 0.5 are discussed as potential trends. All p-values less than 0.1 are provided in text and figures. For clarity in data visualization, baseline values across the two tests (hyperoxia and hypoxia) were averaged and plotted as one value per person at each location where appropriate.

## RESULTS

### Acute mountain sickness

When including all seven timepoints in our linear mixed model and correcting for multiple comparisons, there was not a significant main effect of treatment or timepoint on total morning AMS score. However, the model revealed significant time-by-treatment interactions. There was a highly significant interaction effect at HA1 (b = 3.89, t(92.73) = 4.19, p < .001), indicating that participants in the placebo group experienced a significantly sharper increase in AMS scores at HA1 compared to the ACZ group. A significant interaction was also observed at HA2 (b = 2.22, t(92.43) = 2.44, p = .017), demonstrating a sustained, though slightly smaller, elevation in AMS scores for the placebo group. Interaction effects for all other time points were not statistically significant (**Figure 2**).

**Figure 2.**
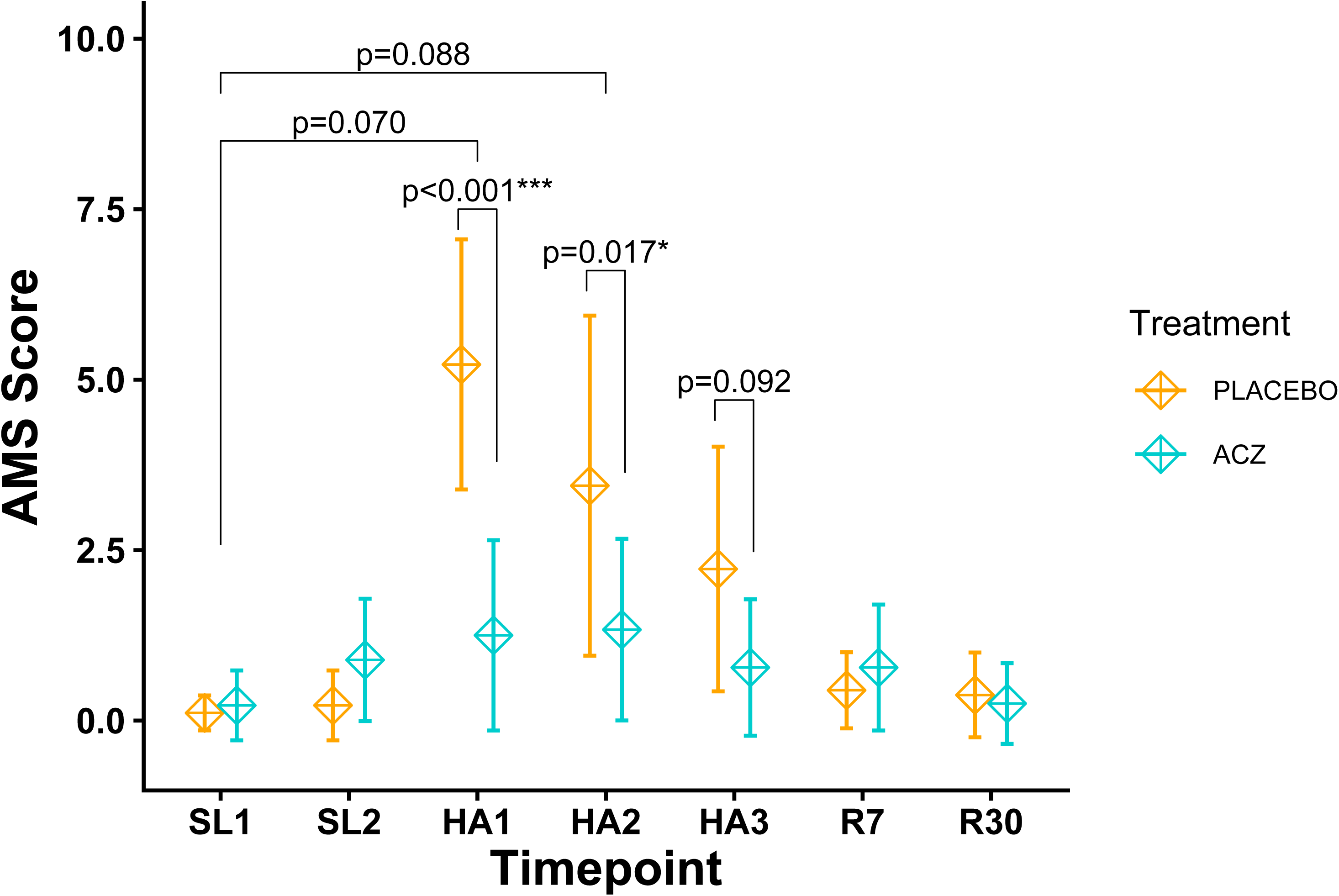
Acute Mountain Sickness (AMS) scores. Mean AMS scores for placebo and acetazolamide (ACZ) groups are provided at each timepoint. Two measures were taken at sea level, prior to ascent: SL1 was collected prior to starting treatment and SL2 was taken on the day of ascent after starting treatments. Measures were then taken each morning at high altitude (HA1, HA2, HA3), as well as 7 (R7) and 30 (R30) days after returning to sea level. Error bars represent 95% confidence intervals.

### Resting physiological parameters

There was a significant main effect of location on resting SpO_2_, demonstrating that resting SpO_2_ was lower at HA compared to SL (b = -12.26, t(16.00) = -11.46, p < .001, **Figure 3A, Figure S1A-C**). However, there was no main effect of treatment. The two-way location-by-treatment interaction showed a marginal trend (b = 2.68, t(16.00) = 1.77, p = .096), suggesting the drop in SpO_2_ at HA may be mitigated by the ACZ treatment compared to the placebo condition. Indeed, a post-hoc test revealed SpO_2_ was marginally significantly higher in the ACZ group compared to the placebo group at HA (estimate = -2.66, t(28.80) = -2.03, p = .051). In a separate analysis comparing the change in SpO_2_ from SL to HA across treatment groups by unpaired t-test, there was no significant difference across treatment groups (p=.099).

**Figure 3.**
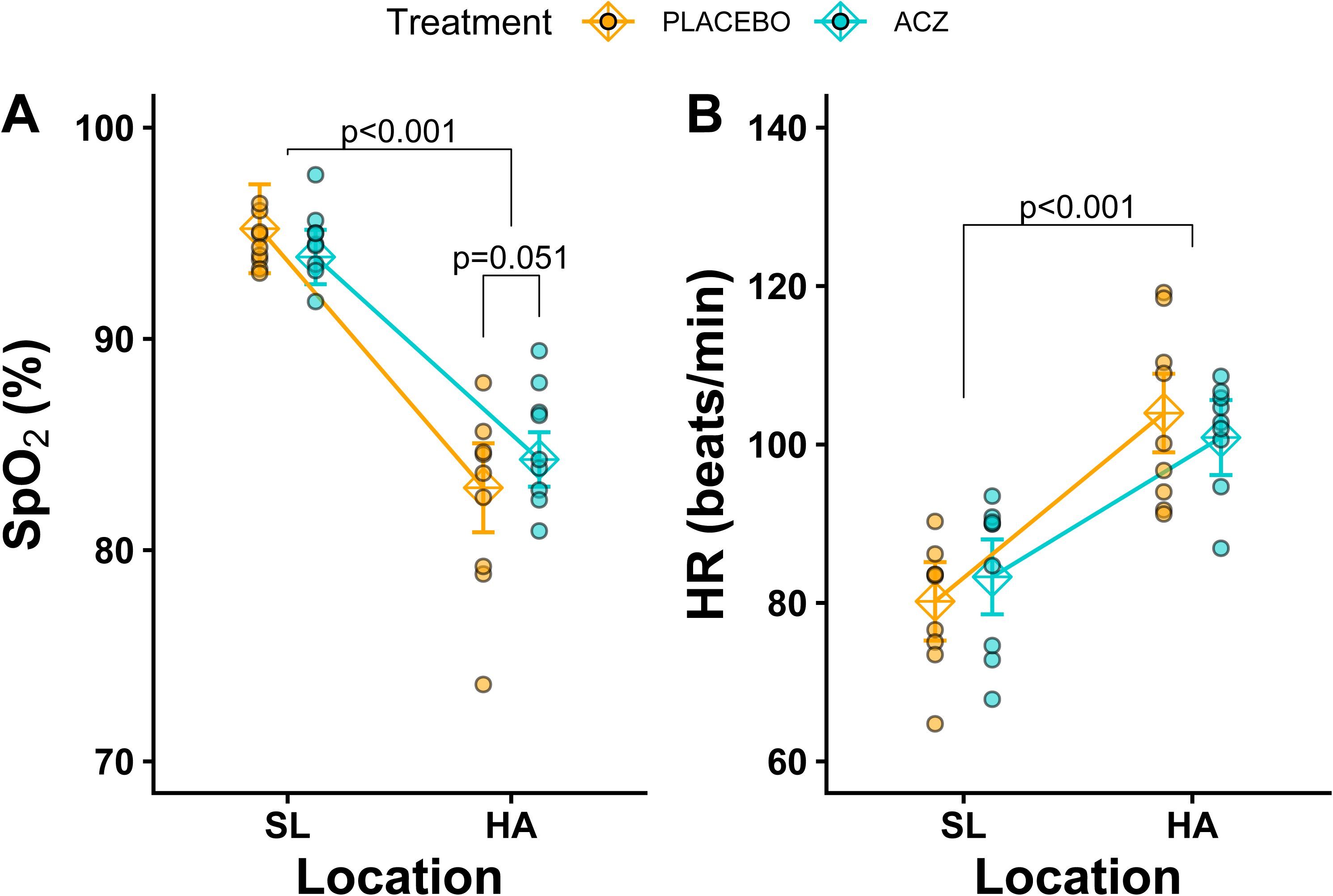
Baseline physiological parameters. Resting HR **(A)** and SpO_2_ **(B)** values are provided for each participant at sea level (SL) and after 2 days at high altitude (HA). Individual datapoints represent the mean of two separate resting measurements taken prior to the hyperoxic and hypoxic chemoreflex trials. Mean values for each group are provided and error bars represent 95% confidence intervals. Significant differences across groups are indicated.

There was a significant main effect of location on resting heart rate, demonstrating that resting heart rate was higher at HA compared to SL to compensate for the reduced SpO_2_ (b = 23.76, t(16.00) = 8.00, p < .001, **Figure 3B, Figure S1D-F**). However, there was no significant main effect of treatment, and the two-way location-by-treatment interaction was not significant, indicating that the increase in resting heart rate experienced at HA did not differ between the placebo and ACZ treatments. In a separate analysis, the change in heart rate from SL to HA did not differ across treatment groups.

There were no significant main effects or interactions between location and treatment on systolic or diastolic blood pressure.

### Baseline breathing

There was a significant main effect of location on resting end-tidal P_CO2_, demonstrating that values were significantly lower at HA compared to SL (b = -8.19, t(16.00) = -8.58, p < .001). The main effect of ACZ treatment was not significant. However, there was a significant two-way location-by-treatment interaction (b = -3.43, t(16.00) = -2.54, p = .022). This interaction indicates that the impact of the treatment on resting end-tidal P_CO2_ differed significantly depending on whether participants were at SL or HA. Indeed, post-hoc tests revealed that at HA, resting end-tidal P_CO2_ was higher in the placebo group compared to the ACZ group (estimate = 5.18, t(23.70) = 3.48, p = .002, **Figure 4A**). In a separate analysis, the change in end-tidal P_CO2_ from SL to HA was significantly different across treatment groups (p = .023).

**Figure 4.**
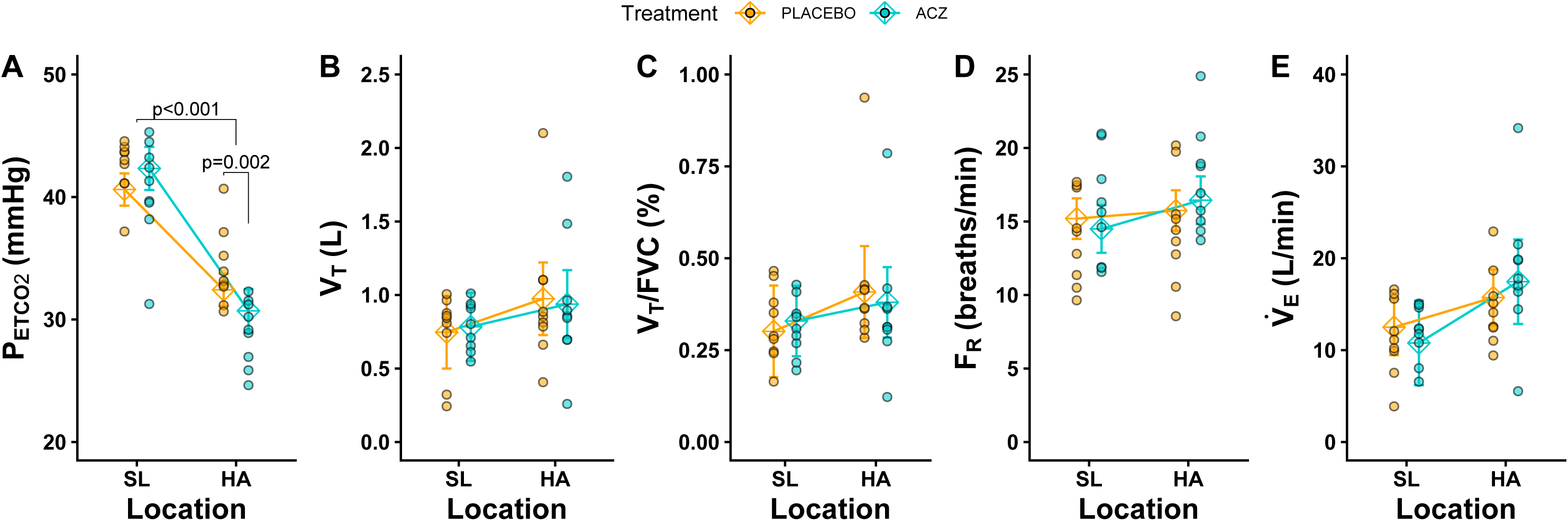
Baseline respiratory parameters. Resting end-tidal P_CO2_ (P_ETCO2_) **(A)**, tidal volume (V_T_) **(B)**, VT as a function of forced vital capacity (FVC) **(C)**, breathing frequency (F_R_) **(D)**, and minute ventilation (V_E_) **(E)** values are provided for each participant at sea level (SL) and after 2 days at high altitude (HA). Individual datapoints represent the mean of two separate resting measurements taken prior to the hyperoxic and hypoxic chemoreflex trials. Mean values for each group are provided, and error bars represent 95% confidence intervals. Significant differences across groups are indicated. Results of linear mixed models are provided at the top of each plot.

Despite these effects on end-tidal P_CO2_, baseline breathing parameters measured prior to ventilatory chemoreflex testing were not impacted by location or treatment. There was no main effect of location or treatment, and no significant two-way interaction, on tidal volume, tidal volume/FVC, breathing frequency, or minute ventilation (**Figure 4B-E, Figure S1D-F**).

### Chemoreflexes

**Figure 1** provides a representative trace of ventilation as a function of end-tidal P_CO2_ under hyperoxic and hypoxic test conditions at SL and HA. We first evaluated the effects of location and treatment on the VRT. There was a significant main effect of location on the VRT, showing that VRTs were significantly lower at HA compared to SL (b = -9.50, t(18.27) = -4.27, p < .001, **Figure 5A**). There was also a significant main effect of test, indicating that VRTs were significantly lower during hypoxic tests compared to hyperoxic tests (b = -3.41, t(18.94) = -2.15, p = .045). In contrast, the main effect of ACZ treatment and all two-and three-way interactions were not statistically significant, indicating that the combined impact of location and testing condition on the VRT was not significantly altered by the administration of ACZ. In separate analyses of the changes of the VRT from SL to HA, there were no significant differences across treatment groups.

**Figure 5.**
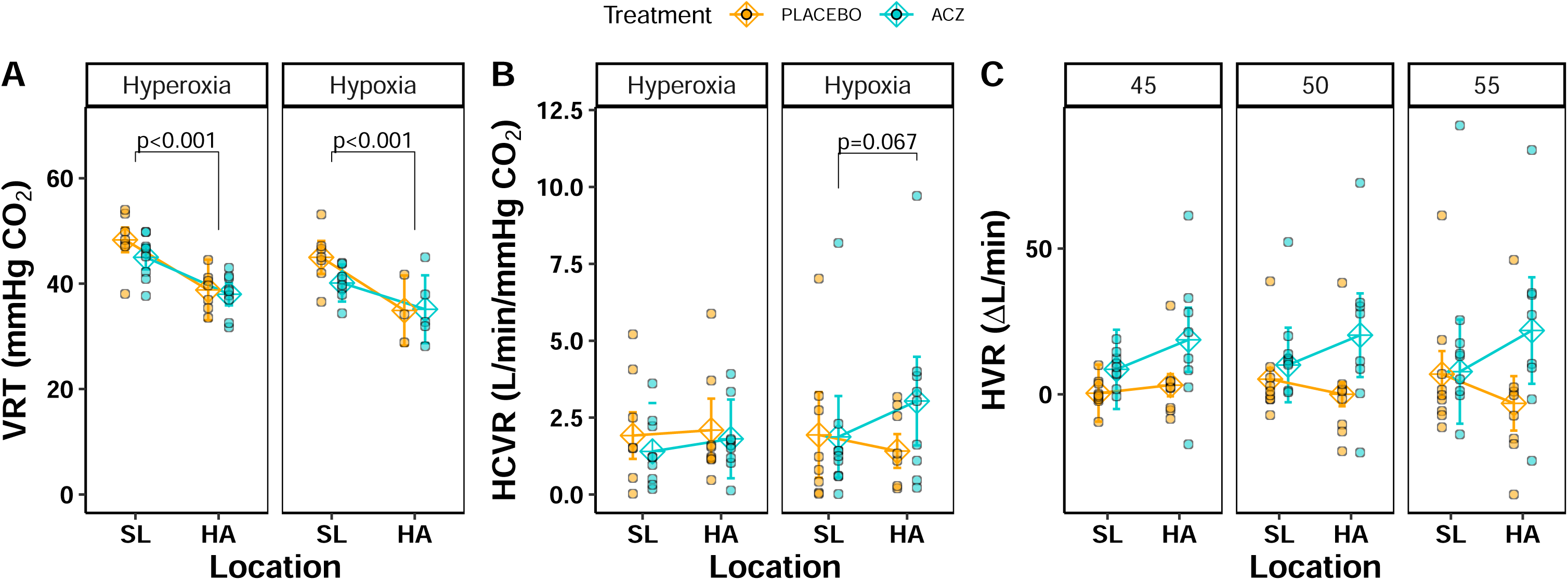
Chemoreflex characteristics. Plots demonstrate ventilatory recruitment thresholds (VRT) **(A)** and hypercapnic ventilatory responses (HCVR) **(B)** collected under hyperoxic and hypoxic conditions, as well as hypoxic ventilatory responses (HVR) collected at three controlled end-tidal P_CO2_ levels (45, 50, and 55 mmHg) **(C)**. Individual values are provided for each participant at sea level (SL) and after 2 days at high altitude (HA). Mean values for each group are provided and error bars represent 95% confidence intervals. Significant differences across groups are indicated.

There were no significant main effects of location, test, or treatment on the HCVR. Similarly, none of the two-way interactions were significant. However, the three-way location-by-treatment-by-test interaction trended toward significance (b = 1.45, t(15.00) = 1.88, p = .079), suggesting a potential effect where the combined impact of HA and hypoxia on the HCVR may differ depending on whether participants received the placebo or ACZ treatment. This may be driven by sensitization to the combined hypoxia and hypercapnia stimulus in the ACZ group at HA, since post-hoc tests revealed a trend toward higher HCVR values in hypoxia at HA in this group only (estimate = -1.16, t(27.50) = -2.60, p = .067, **Figure 5B**). In addition, in separate analyses of the changes of the HCVR from SL to HA, the change in HCVR was significantly elevated at HA during the hypoxia treatment in the ACZ group only (t(13.9) = -2.5, p = 0.027).

Finally, we evaluated the impacts of location, treatment, and P_CO2_ on the HVR. We first examined the HVR values calculated as the change in ventilation between hyperoxic and hypoxic treatments at controlled P_CO2_ levels.^10^ For this measurement, there were no significant main effects of location, treatment, or P_CO2_ (**Figure 5C**). However, there was a highly significant location-by-P_CO2_ interaction (b = -6.78, t(51.31) = -4.50, p < .001). The remaining two-way interactions were not statistically significant. There was also a highly significant three-way location-by-treatment-by-P_CO2_ interaction (b = 8.83, t(50.64) = 4.25, p < .001). This indicates that the linear relationship between P_CO2_ and the HVR at HA varies depending on whether participants received the placebo or ACZ treatment. A post-hoc pairwise comparison was conducted to examine the difference between the treatment groups at HA. This analysis revealed a marginal but insignificant difference between treatment groups (estimate = -31.13, t(15.7) = - 1.94, p = .070), indicating a trend toward lower HVR scores in the placebo group compared to the ACZ group at HA under specific P_CO2_ levels. Within this dataset, there was one significant outlier among the ACZ group with an HVR above Δ100 L/min at HA in hypoxia, resulting from calculations extrapolating steep HCVR slopes in hypoxia. These results persisted regardless of whether this outlier was included or not.

We also examined HVR values as calculated in Duffin (2007), as the change in HCVR slope and VRT between hyperoxic and hypoxic tests.^8^ Similar to the prior calculation method, there were no significant main effects of location or treatment on the ΔHCVR. However, the two-way location-by-treatment interaction showed a trend (b = 1.45, t(15.00) = 1.88, p = .079) suggesting that the relationship between location and the ΔHCVR may tend to differ depending on whether participants received the placebo or ACZ treatment. There were no main effects or interactions between location and treatment on the ΔVRT.

## DISCUSSION

This study revealed that ACZ may influence the chemoreflex control of breathing by modulating the interaction between the peripheral and central components of this reflex. As in prior studies by our group and others, we found that the VRT is reduced with acclimatization to HA, resulting in a higher ventilation rate at a given P_CO2_ (**Figure 1**, **Figure 5A**).^20,28,29^ In studies that use a steady-state method or do not identify a VRT, this characteristic is sometimes referred to as the “HCVR setpoint”. However, in this study, the effect of altitude on the VRT does not seem to be impacted by ACZ. This contrasts with prior studies showing that ACZ alone, tested at SL in the absence of sustained hypoxia exposure, is sufficient to reduce the VRT or HCVR setpoint.^12–14,18,30^ Although some studies find no effect of ACZ on the VRT^11^, this leftward shift in the HCVR curve is well documented under these conditions. However, we could not identify a study that reported the VRT or HCVR setpoint at high altitude after several days of acclimatization and ACZ treatment. It is possible that the impact of high-altitude acclimatization on the VRT, or HCVR setpoint, overshadows underlying effects of ACZ, or that there is a lower limit to the VRT which is reached after several days of acclimatization.

In contrast, we did find a highly significant three-way interaction between treatment, location, and test (hypoxia versus hyperoxia) on the HVR (**Figure 5C**), suggesting the relationship between HVR and the P_CO2_ at which it is measured is significantly altered with ACZ. This was further supported by our findings that the change in HCVR from SL to HA was significantly higher in the ACZ group under hypoxic conditions but not hyperoxic conditions.

These results also contrast prior studies, which report no impact of ACZ on the HVR, or even a decrease in HVR with acute ACZ treatment, likely due to hyperventilation-induced hypocapnia, since the HVR is lower at lower arterial P_CO2_ levels.^14^ However, as with the VRT result, many of these studies measured HVR at SL only.^13,14,18^

Our data likely remains consistent with these prior findings. Our HVR values calculated at 45 mmHg P_CO2_ were comparatively low, and the degree of increase in the HVR at HA is dependent on the P_CO2_ at which it is measured, with higher HVRs measured at higher P_CO2_ values.^9,10^ 45 mmHg falls below the VRT for most participants (**Figure S3**), and therefore HVR values calculated at this level tend to be small but still above zero on average (mean: 4.22 ±7.06 L/min). This is significant because many steady-state HVR protocols, which allow poikilocapnia, or even isocapnia near or slightly above the eupneic P_CO2_, indeed provide small HVR values.

While these may be significant responses under some physiological conditions, the method performed here, as well as the more detailed protocol recently described by Guluzade et al. (2023), may be more powerful for teasing apart treatment effects and individual differences in the HVR by providing more substantial P_CO2_ stimuli and therefore higher and more variable HVR values. However, one consideration when using this method to determine the effects of HA exposure, is how to calculate the HVR when the range of experienced P_CO2_ values during testing may not overlap across HA and SL tests (as demonstrated in **Figure 1** – note data collected at HA does not include data at 55 mmHg P_CO2_, but data collected at SL does).

In addition to these effects on breathing control, we replicate prior findings demonstrating significant improvements in AMS symptoms with ACZ treatment, with dosages ranging from 125 to 500 mg taken twice per day.^31^ Our data supports the efficacy of this lower dosage of ACZ for prophylactic treatment of AMS. This dosage was also sufficient to result in significant reductions in P_CO2_ compared to a control group, as well as increases in daytime SpO_2_ (p=0.051), which likely plays a role in the improved AMS symptoms.^32^

This study may have some limitations. Many of our results indicate trends for significant effects of ACZ treatment. It is possible that our sample size of 18 provided insufficient power to detect some of these effects. In addition, due to logistical constraints, treatment with ACZ was initiated after SL chemoreflex measures were completed, preventing us from determining the impact of this ACZ dose on baseline breathing control.

In conclusion, we found that the current recommended prophylactic use of low-dose ACZ is highly effective at alleviating AMS symptoms during the first few days of high-altitude travel. We also show that this dosage is sufficient to impact the neural control of breathing, as evidenced by reduced end-tidal P_CO2_ likely caused by chronic elevation of alveolar ventilation, as well as sensitizing the interaction between chemoreflex responses to hypoxia and hypercapnia at high altitude.

## ACKNOWLEDGEMENTS

We are grateful to all participants who took part in this study. We are also thankful to the staff at Barcroft Station and the White Mountain Research Station for their hospitality and logistical support. Special thanks to Ani Darakchyan and Ramesh Upadhyayula who provided pharmacy services for this study.

## AUTHOR CONTRIBUTIONS

ECH conceptualized the work. Data was curated by ECH, HP, AV, VP, DA, and SL. Formal analysis was conducted by ECH. Funding was acquired by ECH and AV. HB contributed to the study methodology and project administration.

## STATEMENTS AND DECLARATIONS

### Ethical considerations

This study was approved by the University of California Riverside Institutional Review Board (protocol #30687). The study was conducted in accordance with the *Declaration of Helsinki*, including registration in a database (clinicaltrials.gov NCT07517068).

### Consent to participate

A digital version of the complete consent form was provided to all participants prior to their first study visit to review on their own. During their first study visit, we reviewed the written consent form with participants, provided them with time to ask any questions, and each participant provided written, informed consent in their native language (English).

### Consent for publication

Not applicable.

### Declaration of conflicting interest

The authors declared no potential conflicts of interest with respect to the research, authorship, and/or publication of this article

### Funding statement

This work was supported by a Herbert N. Hultgren Grant from the Wilderness Medical Society to ECH and a White Mountain Mini Grant to AV.

### Data availability

Raw data used in the analyses reported here are available as supplemental material.

## Notes

### Competing Interest Statement

The authors have declared no competing interest.

### Clinical Trial

NCT07517068

### Author Declarations

The Institutional Review Board of the University of California, Riverside gave ethical approval for this work.

