## Supplementary figures and images for "Effects of Prophylactic Low-Dose Acetazolamide on the Chemoreflex Control of Breathing and Acute Mountain Sickness at High Altitude"

### Supplemental Figure 1

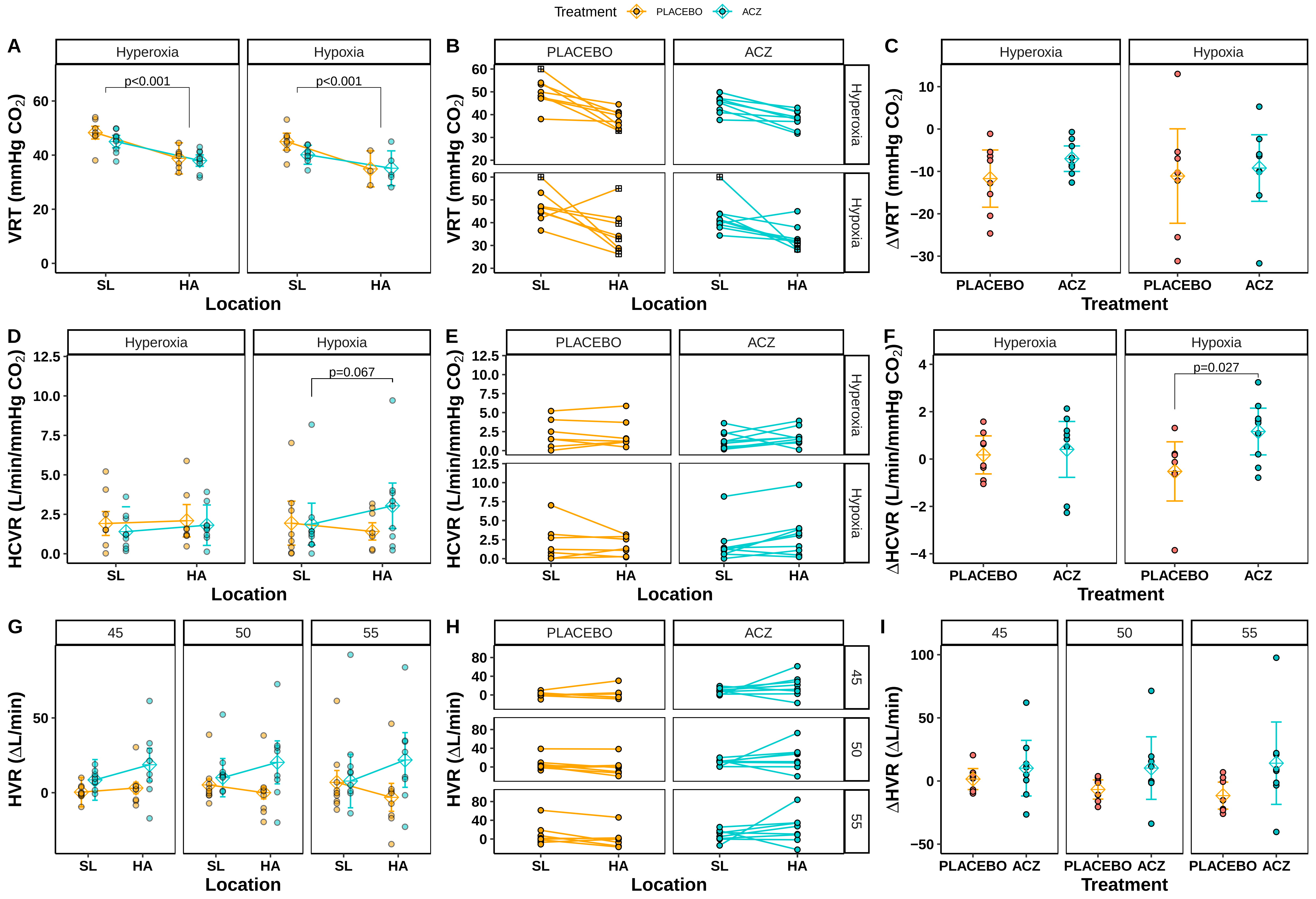

### Supplemental Figure 2

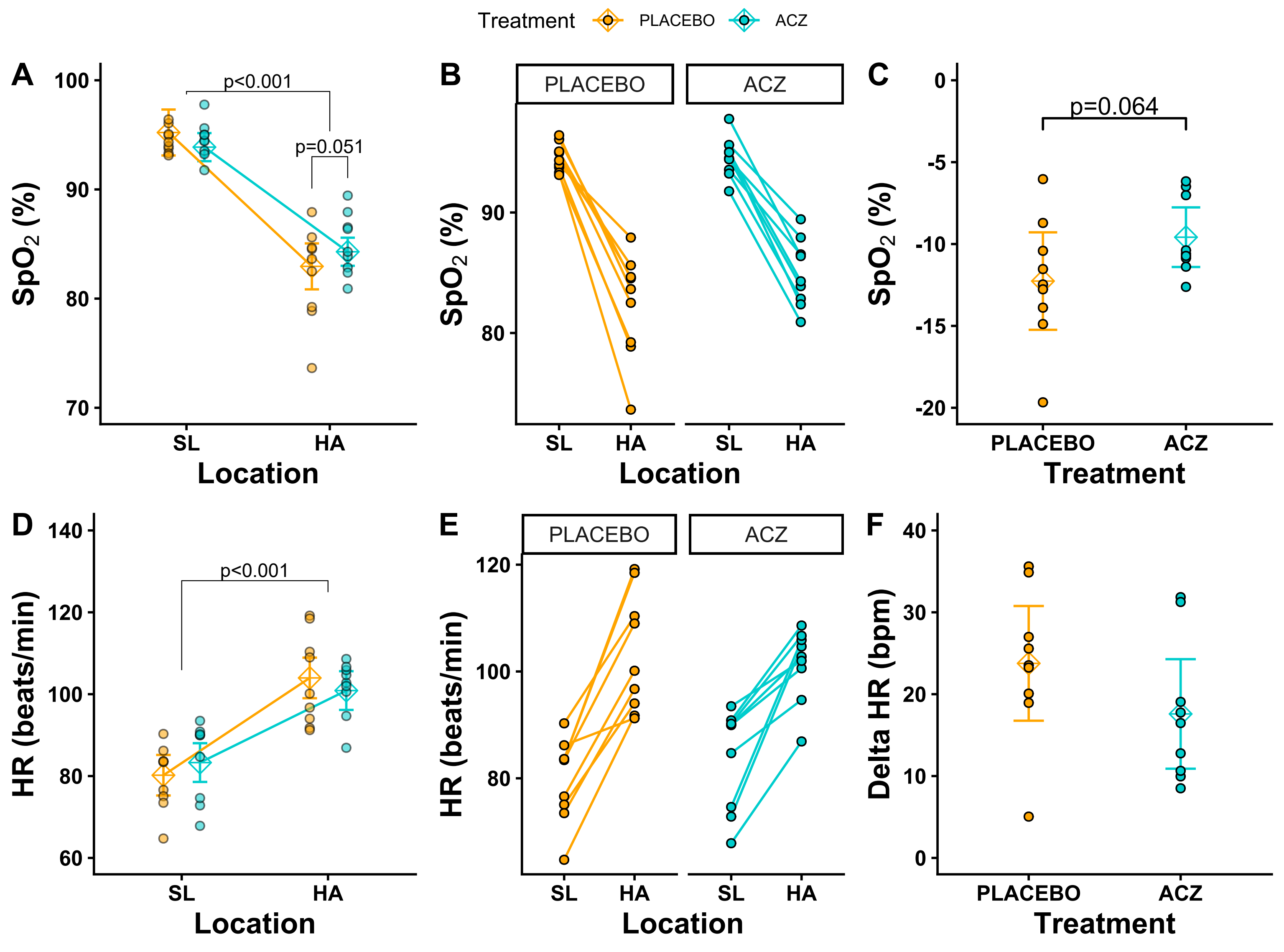

### Supplemental Figure 3

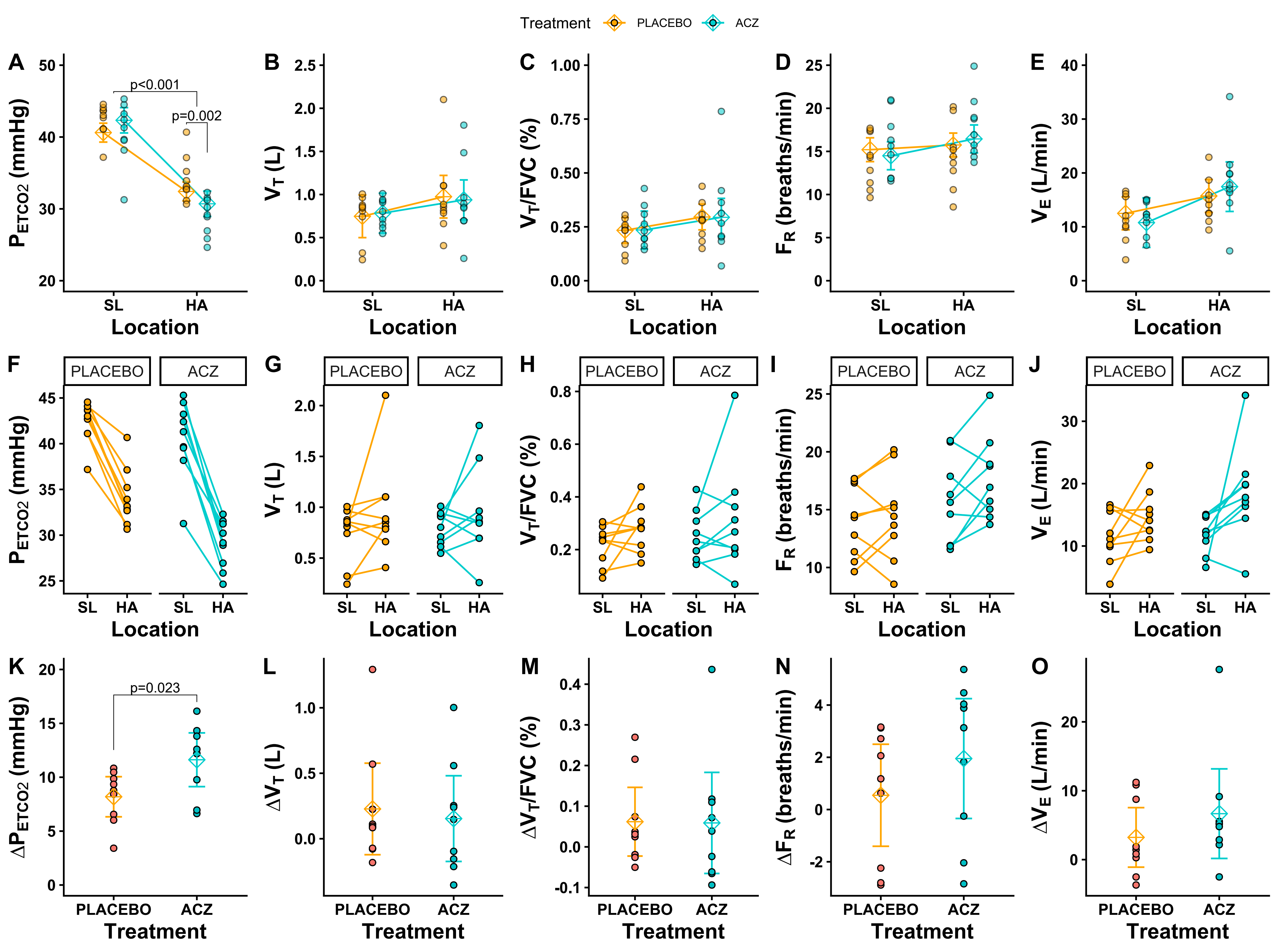

### Supplemental Figure 4

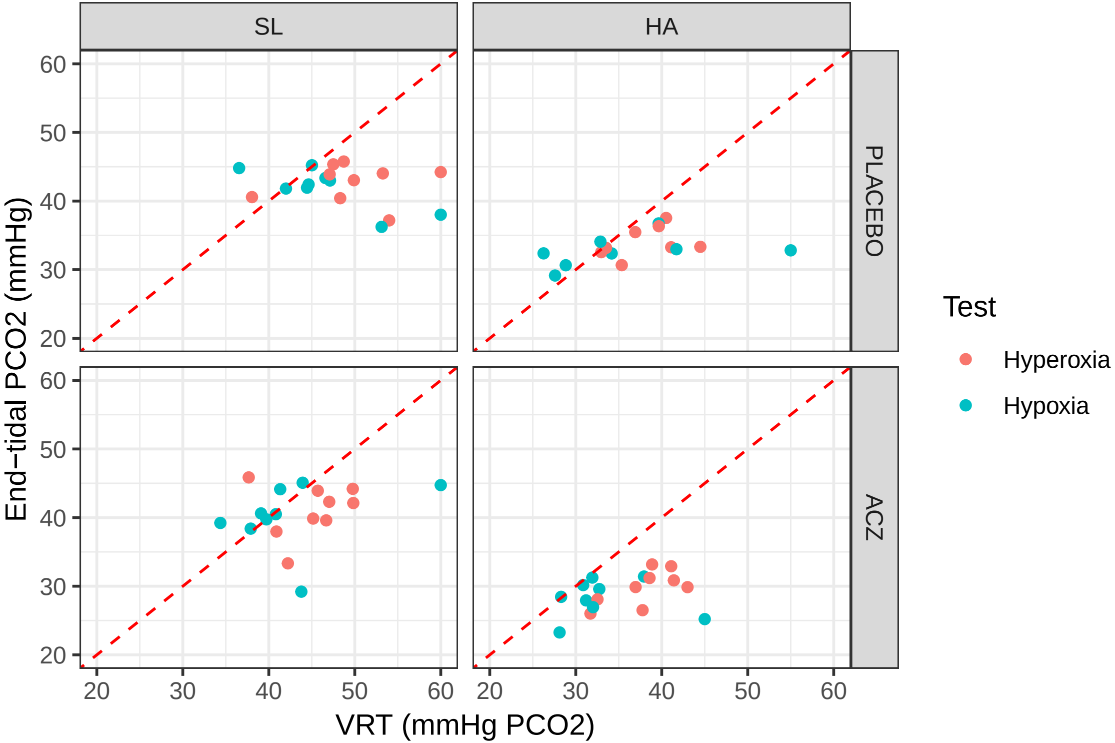
